# Targeted Proteomic Biomarker Profiling Using NULISA in a cohort enriched with risk for Alzheimer’s Disease and Related Dementias

**DOI:** 10.1101/2024.11.28.24318162

**Authors:** Ramiro Eduardo Rea Reyes, Rachael E. Wilson, Rebecca E. Langhough, Rachel L. Studer, Erin M. Jonaitis, Julie E. Oomens, Elizabeth M. Planalp, Barbara B. Bendlin, Nathaniel A. Chin, Sanjay Asthana, Henrik Zetterberg, Sterling C Johnson

## Abstract

**INTRODUCTION:** Targeted proteomic assays may be useful for diagnosing and staging Alzheimer’s disease and related dementias (ADRD). We evaluated the performance of a 120-marker central nervous system (CNS) NUcleic acid-Linked Immuno-Sandwich Assay (NULISA) panel in samples spanning the AD spectrum.

**METHODS:** Cross-sectional plasma samples (n=252) were analyzed using Alamar’s NULISAseq CNS panel. ROC analyses demonstrated NULISAseq-pTau217 accuracy in detecting amyloid (A) and tau (T) PET positivity. Differentially expressed proteins were identified using volcano plots.

**RESULTS:** NULISAseq-pTau217 accurately classified A/T PET status with ROC AUCs of 0.92/0.86. pTau217 was upregulated in A+, T+, and impaired groups with log2-fold changes of 1.21, 0.57 and 4.63, respectively, compared to A-. Interestingly, pTDP43-409 was also upregulated in the impaired group and correlated with declining hippocampal volume and cognitive trajectories.

**DISCUSSION:** This study shows the potential of a targeted proteomics panel for characterizing brain changes pertinent to ADRD. The promising pTDP43-409 findings require further replication.

## Background

The pathological hallmarks of Alzheimer’s disease (AD) include amyloid-beta plaques and neurofibrillary tau tangles. Abnormal accumulation of amyloid (A) and tau (T) are used to detect and stage AD [1] and can be ascertained from cerebrospinal fluid (CSF) markers, positron emission tomography (PET) and more recently, plasma markers [2], which are fast increasing in accuracy, sensitivity, breadth, and scalability [3]. Simultaneously, there is an urgent need to detect and stage other neurodegenerative diseases that may be contributing to cognitive impairment and dementia along with or instead of AD. A broad biomarker panel that is capable of detecting AD and non-AD proteinopathies will be especially informative since it is now acknowledged that co-pathology with AD is rather common, particularly alpha-synuclein [4] and TDP-43 pathologies [5, 6].

Recent advancements in proximity ligation assays have made it possible to measure large panels (>100) of central nervous system (CNS)-related proteins in low sample volumes [7]. These assays open the door to both biological characterization of clinical phenotypes and broad exploration of novel biomarker candidates, all within a single analysis. Recognizing this, we evaluated the performance and potential utility of the Nucleic Acid-Linked Immuno-Sandwich Assay CNS panel with detection by next generation sequencing (NULISAseq) for characterizing AD pathology and identifying candidate biomarkers of non-AD proteinopathies in plasma samples spanning the AD spectrum.

We selected 252 cross-sectional plasma samples from well-characterized participants enrolled in either the Wisconsin Registry for Alzheimer’s Prevention (WRAP) or the Wisconsin Alzheimer’s Disease Research Center (WADRC) clinical core cohorts. The sample selection was based on the availability of amyloid and tau PET status and clinical diagnosis. Our initial aim was to determine if the NULISAseq CNS panel accurately detects amyloid and tau pathologies using tau phosphorylated at threonine 217 (pTau217), a Core 1 marker for AD diagnosis [1], and to compare pTau217 performance as part of a multiplex panel to the well-established Single molecule array (Simoa) singleplex assay. We then examined the differential expression of the entire panel of biomarkers across known statuses determined by amyloid PET, tau PET, the Amprion seed aggregation assay (synSAA) for oligomeric alpha-synuclein, and clinical diagnosis in order to identify potential markers from the CNS panel for further follow-up. Based on these findings, we explored associations between plasma measurements of TDP-43 phosphorylated at serine 409 (pTDP43-409), a protein related to limbic associated TDP-43 encephalopathy (LATE), with longitudinal changes in cognitive scores and hippocampal volume.

## Methods

### Cohort

Samples were obtained from participants in the WRAP and WADRC studies. WRAP is a longitudinal observational study focused on preclinical AD with 2823 late-middle age adults enrolled at the time of these analyses. Most participants have normal baseline cognition (86.4% cognitively unimpaired (CU); 6.2% mild cognitive impairment (MCI) at enrollment), and the cohort is enriched for parental history of AD. Enrollment in the ADRC is open to participants across the cognitive status continuum and is also enriched for parental history of AD. Participants return for extensive cognitive testing and blood collection approximately every 2 years in WRAP or every 1-2 years in the ADRC, depending on age and cognitive status; participants also have the opportunity to undergo additional biomarker testing, including PET, MRI and lumbar puncture. The WRAP and WADRC studies and associated biomarker testing have been approved by the Institutional Review Board at the University of Wisconsin – Madison and were conducted in accordance with the Declaration of Helsinki. All participants provided informed consent.

### Sample collection and selection

Plasma samples included in this study (n = 252) were collected between 2017-2024 using lavender top K_2_EDTA tubes. The tubes were inverted ten times immediately after collection and again prior to centrifugation 2000 x g for 10 minutes. Plasma was then aliquoted into cryovials for long-term storage at -80 °C.

Sample selection was based on the availability of amyloid (A) and tau (T) PET status within one year of the blood draw (n = 239; PET methods detailed subsequently) with strategic oversampling of those who were PET A+. To improve the diversity of the sample set, we included samples from thirteen additional participants who lacked both amyloid and tau PET imaging but had existing Simoa pTau217 plasma results available to approximate A and T status(see Table 1 for details).

**Table 1.** Sample characteristics across analyses. Percentages are calculated column wise.

| Variable | Overall | CU | MCI | Dementia | *p value* |
| --- | --- | --- | --- | --- | --- |
| N (%) | 252 | 207 (82.1%) | 36 (14.3%) | 9 (3.6%) |  |
| Female, n (%) | 172 (68.3%) | 147 (71.0%) | 19 (52.8%) | 6 (66.7%) | = .095 |
| Age at blood draw, mean (SD) | 70.1 (7.1) | 69.2 (7.0) | 73.8 (6.2) | 76.1 (6.2) | < .001 |
| ***Race composition, n (%)*** |  |  |  |  |  |
| American Indian or Alaskan Native |  | 9 (3.6%) |  |  |  |
| Black or African American |  | 27 (10.7%) |  |  |  |
| White |  | 214 (84.9%) |  |  |  |
| Other |  | 2 (0.8%) |  |  |  |
| **Stratified by PET VR status** |  |  |  |  |  |
| Variable | Overall | A-/T- | A+/T- | A+/T+ | *p value* |
| N (%) | 239 | 74 (31.0%) | 90 (37.7%) | 75 (31.4%) |  |
| Female, n (%) | 162 (67.8%) | 52 (70.3%) | 59 (65.6%) | 51 (68.0%) | = .812 |
| CU, n (%) | 195 (81.6%) | 72 (97.3%) | 88 (97.8%) | 35 (46.7%) | < .001 |
| Age at blood draw, mean (SD) | 70.4 (7.0) | 67.9 (8.0) | 70.6 (6.0) | 72.5 (6.5) | < .001 |
| BA or more education, n (%) | 168 (70.3%) | 52 (70.3%) | 65 (72.2%) | 51 (68.0%) | = .840 |
| Years of PACC3 follow-up, mean (SD) | 9.9 (4.7) | 10.0 (3.9) | 10.6 (4.5) | 8.8 (5.3) | = .042 |
| Practice, median [Q1, Q3] | 5.00 [4.00, 6.00] | 5.00 [4.00, 6.00] | 6.00 [5.00, 6.00] | 5.00 [4.00, 6.00] | = .074 |
| Years of volumetric follow-up, mean (SD) | 7.1 (4.9) | 7.5 (4.7) | 7.6 (4.6) | 6.0 (5.2) | = .062 |
| Volumetric visits, median [Q1, Q3] | 4.00 [2.00, 6.00] | 4.50 [2.00, 6.00] | 4.00 [2.00, 5.00] | 3.00 [1.00, 5.00] | = .095 |
| **Stratified by synSAA status** |  |  |  |  |  |
| Variable | Overall | SynSAA- | SynSAA+ | *p value* |  |
| N (%) | 108 | 91 (84.3%) | 17 (15.7%) |  |  |
| Female, n (%) | 69 (63.9%) | 60 (65.9%) | 9 (52.9%) | = .454 |  |
| CU, n (%) | 88 (81.5%) | 76 (83.5%) | 12 (70.6%) | = .358 |  |

### Sample analysis

Samples were analyzed at the University of Wisconsin – Madison ADRC Biomarker Lab using the NULISAseq CNS kit (Alamar Part# 800104, Lot# 2407305) according to manufacturer instructions. Briefly, samples were thawed at room temperature, centrifuged for 5 minutes at 10,000 x g and 4 °C, and transferred to the 96-well sample plate provided with the reagent kit.

The plate was sealed and spun for 1 minute at 1000 x g and 4 °C. Utilizing the automated Alamar Argo HT workflow, target analytes within each sample were bound through multiple capture, wash, and release steps, resulting in immunocomplexes with both sample-specific and target-specific DNA tags that were pooled into a single library for next generation sequencing [7]. The library was sequenced using a P2 100 cycle flow cell (Illumina Part# 20046811, Reagent Lot# 20841078, Flow cell Lot# 20846922) on a NexSeq1000. After sequencing, the FASTQ file was uploaded to Argo Command Center for automatic normalization and conversion to NULISA protein quantification (NPQ) units for each biomarker. Samples were analyzed in three batches with eighty-four samples per batch. A pooled plasma control sample (prepared in-house) was analyzed at the start and end of the plate to assess within-plate (n = 2) and between-plate (n = 6) reproducibility. The within-plate percent coefficient of variation (%CV) ranged from 0.01% to 7.5% for most biomarkers. The exception was for hemoglobin subunit 1 (HBA1), which ranged from 10.8% to 31.8%CV. For each of the three batches, 90% of targets had a within-plate CV ≤ 2.0%. The between-plate %CV was similar, ranging from 0.29% to 5.6%CV, except for HBA1 (19.1%CV).

### Neuropsychological assessment and cognitive status

WRAP and ADRC participants completed comprehensive cognitive batteries at each visit, assessing memory, executive function, language, and other cognitive abilities, along with self-reported and study partner assessments of daily functioning [8–10]. Scores from individual tests were cross-walked where needed for consistency across cohorts and over time[10, 11].

Cognitive status was determined through a multidisciplinary consensus review in each cohort [10, 12], using validated diagnostic criteria for MCI [13] and dementia [14]. Consensus review and cognitive status determination were blind to any biomarker information for both studies.

For this study, we used two composite cognitive scores: a modified, three-test version of the Preclinical Alzheimer’s Cognitive Composite (PACC3-TMTB; incorporating Rey AVLT total [15], Logical Memory A Delayed [16]; Trail-Making Test B [17]), which is sensitive to preclinical Alzheimer’s decline [10, 18, 19]; and a memory composite (Rey AVLT Immediate & Delayed; Logical Memory A Immediate & Delayed). The scores from each subtest were rescaled such that first observations in cognitively unimpaired individuals were distributed ∼*N*(0,1) with higher scores representing better performance [10]; an unweighted average of component scores was computed, and the result similarly rescaled to produce each composite.

### Imaging methods

As noted, most participants in this study (n = 239) had completed at least one amyloid PET scan and one tau PET scan within a year of the plasma sample selected for NULISA analysis. The methods for acquiring [C-11]PiB PET (amyloid) and [F-18]Florquinitau (FQT; also known as MK-6240 for tau) PET imaging, the corresponding T1-weighted MRI procedures, and described the visual evaluation of amyloid and tau PET positivity were previously have been reported previously [20]. Briefly, for amyloid PET positivity, raters assigned a positive label only in cases of substantial cortical gray matter signal in one or more of medial, lateral, superior, or ventral lobe segments from the parietal, temporal, frontal, or occipital lobes (unilateral or bilateral) [21]. Tau PET positivity required positive signal in both the medial temporal lobe and neocortex. MRI T1-weighted volumes were segmented into gray matter, white matter, and cerebrospinal fluid using SPM12 which was used to derive measures of total brain and intracranial volume. Hippocampal volume was derived using FSL First [22]. All MRI outputs were visually checked for quality control. In the subset of all participants with amyloid and tau scans, samples were selected for NULISA analysis from three groups: A-/T-, n = 74 (31%); A+/T-, n = 90 (37.7%); and A+/T+ n = 75 (31.4%).

### Other fluid biomarkers

Additional fluid biomarker data for the selected samples were included in these analyses for comparison with the NULISAseq CNS panel. We specifically aimed to assess the performance of the pTau217 assay within the CNS panel against the Quanterix Simoa platform, both utilizing the ALZpath antibody. Of the 252 participants in our sample, 218 (86.5%) had been previously analyzed with the ALZpath Simoa pTau217 assay (Quanterix Part# 104570, Lot# 999008) on a Quanterix HD-X according to kit instructions. Additionally, to evaluate the NULISAseq CNS panel’s performance in detecting alpha-synuclein-related pathology in plasma, we compared biomarker expression in participants with evidence of oligomeric alpha-synuclein (Oligo-SNCA), as determined in CSF by the Amprion alpha-synuclein seed amplification assay (synSAA) [23, 24]. Our sample included a subset of 109 participants with available synSAA results [25], 17 (15.6%) of whom were synSAA+ and 92 (84.4%) synSAA-.

### Statistical Analyses

All analyses were performed using R 4.4.0[26] and the R-packages pROC [27], lme4 [28], and emmeans [29] to perform receiver operating characteristic (ROC) analyses, linear mixed models, and post-hoc comparisons, respectively.

### NULISA pTau217 performance

To examine how the NULISAseq CNS disease panel compared to Simoa assays to characterize A and T PET status, we selected the subset of participants who had both Simoa and NULISA data for a plasma sample and who had a PET scan no more than one year from the blood draw (n = 218). We compared the ROC AUC from these platforms by performing separate DeLong’s tests for ROC analyses relative to A PET status and T PET status. For each ROC analysis, we also identified the Youden’s J index threshold.

### Proteomics

To evaluate differential expression of the entire NULISAseq CNS panel, we constructed volcano plots based on linear models with the analyte NPQ concentrations as the dependent variable, and the binary grouping as predictor (analyte NPQ ∼ group) for various groups of interest, specifically A+ vs A-; A+/T- vs A+/T+; all pairwise contrasts of CU, MCI, and dementia; and synSAA- vs synSAA+. We applied FDR correction to the results for each pair of groups, to keep the type I error at the 0.05 level. We used a log_2_ fold change (log_2_ FC) of 0.5 as criterion to determine effects of interest, and an FDR corrected p-value < 0.05. Given the small sample size of the subset with synSAA status, we also included planned comparisons of two alpha-synuclein plasma biomarkers on the NULISA panel, oligomeric alpha-synuclein (Oligo-SNCA) and monomeric alpha-synuclein (Mono-SNCA).

### Exploratory longitudinal analysis

To explore the utility of the NULISAseq CNS panel for detecting other neurodegenerative diseases for which no established fluid biomarkers exist, we examined the relationship between pTDP43-409, a candidate marker for TDP43-associated proteinopathy, and clinical indicators of LATE [30]. Since we do not have gold standard biomarkers for LATE in this cohort, we binarized the group based on pTDP43-409 measurements. To do this, we calculated the mean and SD from the subset of young (less than 65 years old) cognitively unimpaired and amyloid negative participants in our sample (n = 26, mean = 9.83, SD = 0.44), and we used this to scale all the pTDP43-409 values in the full sample. Scaled scores >2 (i.e., more than 2 SD above the CU mean) were categorized as Elevated (presumed higher risk), and the rest as Non-Elevated.

We employed linear mixed-effects models to examine the longitudinal trajectories of the two cognitive composite outcomes (PACC3-TMTB composite and memory composite, n = 234) and hippocampal volume (n = 190). These models included both linear and quadratic age predictors (centered at age 65) and a three-way interaction between PET A/T status (A-/T-, A+/T, A+/T+), pTDP43-409 status, and age, with random intercepts per participant. For the cognitive composite analyses, we included years of education, sex, and practice (number of prior test exposures) as covariates. For the hippocampal volume model, overall brain volume was included as a covariate to determine if participants with elevated pTDP43-409 showed a more drastic hippocampal decline, even after accounting for overall brain atrophy [30]. We removed any non-significant interactions and reported the final models in Table 2.

**Table 2.**
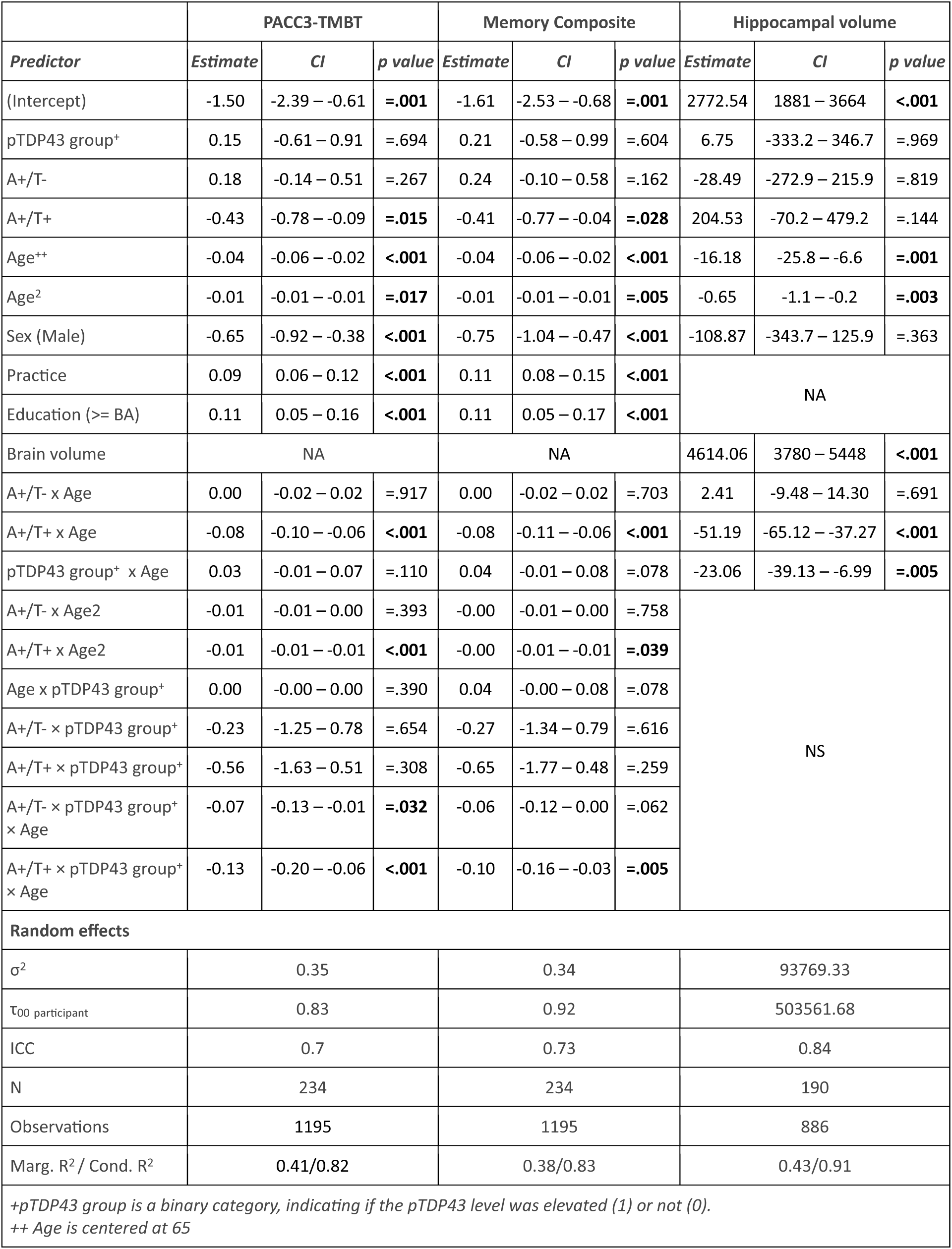
Model output for mixed models.

|  | PACC3-TMBT |  |  | Memory Composite |  |  | Hippocampal volume |  |  |
| --- | --- | --- | --- | --- | --- | --- | --- | --- | --- |
| Predictor | Estimate | CI | p value | Estimate | CI | p value | Estimate | CI | p value |
| (Intercept) | -1.50 | -2.39 – -0.61 | =.001 | -1.61 | -2.53 – -0.68 | =.001 | 2772.54 | 1881 – 3664 | <.001 |
| pTDP43 group<sup>+</sup> | 0.15 | -0.61 – 0.91 | =.694 | 0.21 | -0.58 – 0.99 | =.604 | 6.75 | -333.2 – 346.7 | =.969 |
| A+/T- | 0.18 | -0.14 – 0.51 | =.267 | 0.24 | -0.10 – 0.58 | =.162 | -28.49 | -272.9 – 215.9 | =.819 |
| A+/T+ | -0.43 | -0.78 – -0.09 | =.015 | -0.41 | -0.77 – -0.04 | =.028 | 204.53 | -70.2 – 479.2 | =.144 |
| Age<sup>++</sup> | -0.04 | -0.06 – -0.02 | <.001 | -0.04 | -0.06 – -0.02 | <.001 | -16.18 | -25.8 – -6.6 | =.001 |
| Age<sup>2</sup> | -0.01 | -0.01 – -0.01 | =.017 | -0.01 | -0.01 – -0.01 | =.005 | -0.65 | -1.1 – -0.2 | =.003 |
| Sex (Male) | -0.65 | -0.92 – -0.38 | <.001 | -0.75 | -1.04 – -0.47 | <.001 | -108.87 | -343.7 – 125.9 | =.363 |
| Practice | 0.09 | 0.06 – 0.12 | <.001 | 0.11 | 0.08 – 0.15 | <.001 | NA |  |  |
| Education (>= BA) | 0.11 | 0.05 – 0.16 | <.001 | 0.11 | 0.05 – 0.17 | <.001 |  |  |  |
| Brain volume | NA |  |  | NA |  |  | 4614.06 | 3780 – 5448 | <.001 |
| A+/T- x Age | 0.00 | -0.02 – 0.02 | =.917 | 0.00 | -0.02 – 0.02 | =.703 | 2.41 | -9.48 – 14.30 | =.691 |
| A+/T+ x Age | -0.08 | -0.10 – -0.06 | <.001 | -0.08 | -0.11 – -0.06 | <.001 | -51.19 | -65.12 – -37.27 | <.001 |
| pTDP43 group<sup>+</sup> x Age | 0.03 | -0.01 – 0.07 | =.110 | 0.04 | -0.01 – 0.08 | =.078 | -23.06 | -39.13 – -6.99 | =.005 |
| A+/T- x Age<sup>2</sup> | -0.01 | -0.01 – 0.00 | =.393 | -0.00 | -0.01 – 0.00 | =.758 | NS |  |  |
| A+/T+ x Age<sup>2</sup> | -0.01 | -0.01 – -0.01 | <.001 | -0.00 | -0.01 – -0.01 | =.039 |  |  |  |
| Age x pTDP43 group<sup>+</sup> | 0.00 | -0.00 – 0.00 | =.390 | 0.04 | -0.00 – 0.08 | =.078 |  |  |  |
| A+/T- x pTDP43 group<sup>+</sup> | -0.23 | -1.25 – 0.78 | =.654 | -0.27 | -1.34 – 0.79 | =.616 |  |  |  |
| A+/T+ x pTDP43 group<sup>+</sup> | -0.56 | -1.63 – 0.51 | =.308 | -0.65 | -1.77 – 0.48 | =.259 |  |  |  |
| A+/T- x pTDP43 group<sup>+</sup> x Age | -0.07 | -0.13 – -0.01 | =.032 | -0.06 | -0.12 – 0.00 | =.062 |  |  |  |
| A+/T+ x pTDP43 group<sup>+</sup> x Age | -0.13 | -0.20 – -0.06 | <.001 | -0.10 | -0.16 – -0.03 | =.005 |  |  |  |
| Random effects |  |  |  |  |  |  |  |  |  |
| σ<sup>2</sup> | 0.35 |  |  | 0.34 |  |  | 93769.33 |  |  |
| τ<sub>00</sub> participant | 0.83 |  |  | 0.92 |  |  | 503561.68 |  |  |
| ICC | 0.7 |  |  | 0.73 |  |  | 0.84 |  |  |
| N | 234 |  |  | 234 |  |  | 190 |  |  |
| Observations | 1195 |  |  | 1195 |  |  | 886 |  |  |
| Marg. R<sup>2</sup> / Cond. R<sup>2</sup> | 0.41/0.82 |  |  | 0.38/0.83 |  |  | 0.43/0.91 |  |  |
| +pTDP43 group is a binary category, indicating if the pTDP43 level was elevated (1) or not (0).<br>++ Age is centered at 65 |  |  |  |  |  |  |  |  |  |

## Results

From the 252 samples, 207 (82.1%) were cognitively unimpaired (CU), 36 (14.3%) were MCI, and 9 (3.6%) had dementia at the visit corresponding to the plasma sample. Stratifying the 239 participants with amyloid and tau PET status, 72(30.1%) were A-T-CU, 85(35.6%) were A+T-CU, 33(13.8%) were A+T+ CU. Forty of the forty-five participants with cognitive impairment (MCI or dementia, n=45) were A+T+. Additional participant characteristics are shown in Table 1.

### NULISAseq CNS pTau217 performance

Relative concentrations obtained from the pTau217-NULISA(2^NPQ^) correlated well with absolute concentrations obtained from the pTau217-Simoa assay (pg/mL) (n=218, Figure 1A; *r* = 0.84, *p* < .001). ROC analyses using each assay to classify participants by amyloid PET positivity showed similarly robust performance for each (pTau217-NULISA: AUC=0.92 (0.88-0.97); pTau217-Simoa: AUC=0.91 (0.86-0.95; DeLong z=0.88, p=0.378; Figure 1B). For classification by tau PET positivity, AUCs for both assays were within an acceptable range (>.80) [31], but the pTau217-NULISA AUC was slightly lower than that for the pTau217-Simoa assay (pTau217-NULISA: AUC=0.86 (0.81-0.90); pTau217-Simoa: AUC=0.89 (0.85-0.94); DeLong z=-2.19, p=.028) (Figure 1C).

**Figure 1.**
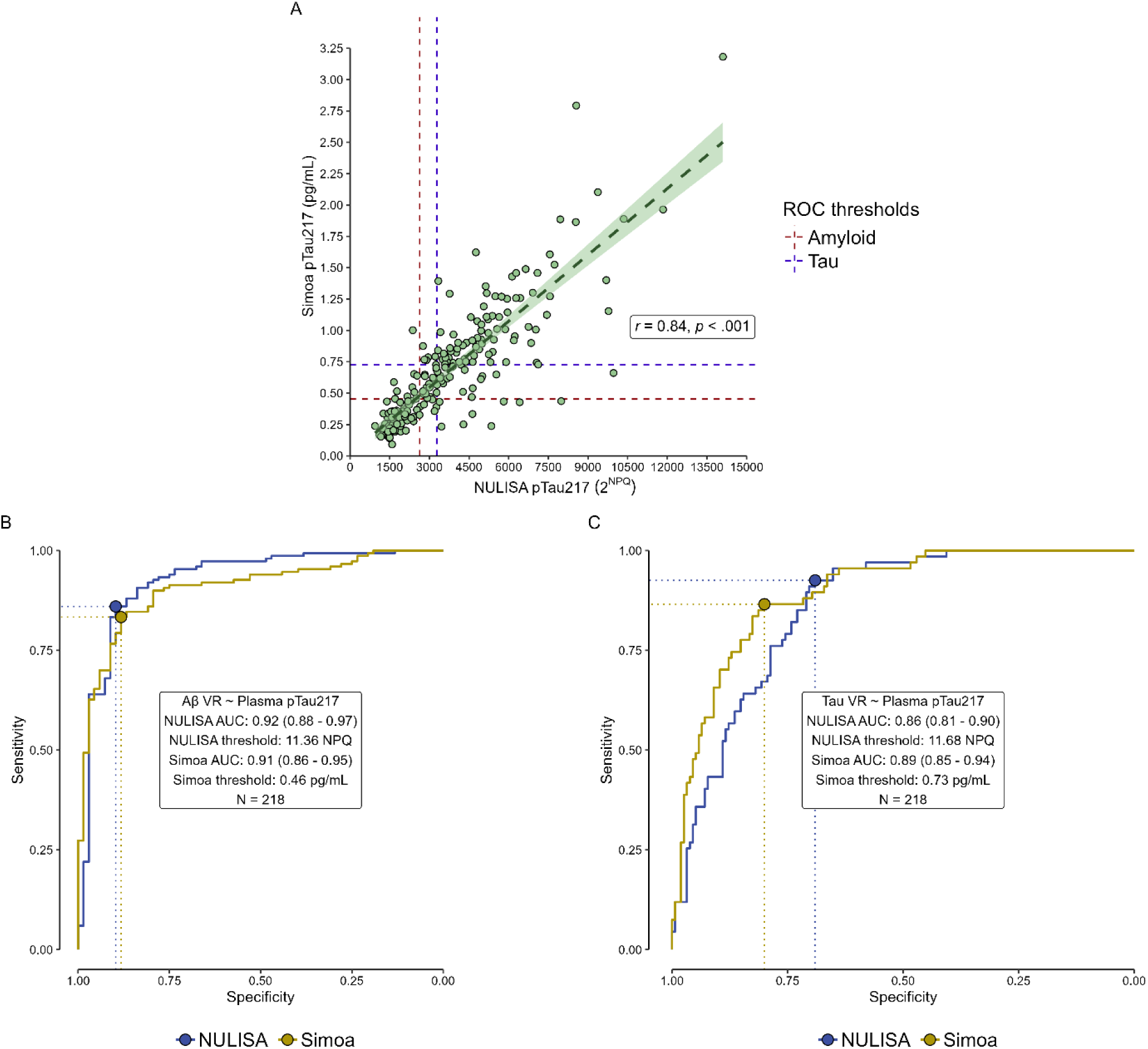
Performance comparison between NULISA and Simoa for subset of n=218 with both plasma assays and amyloid PET visual read. A) Correlation between NULISA 2NPQ and Simoa pg/mL concentrations. Dashed lines indicate the Youden’s cut-off for the NULISA (vertical line) and Simoa (horizontal line) assays, derived from their respective ROCs for amyloid visual read (red) and tau visual read (purple). B-C) ROC curves from NULISA (blue) and Simoa (yellow), based on amyloid visual read (B) and tau visual read (C).

### Differential protein expression

The volcano plots in Figure 2 illustrate results of the linear model fits predicting NPQ for each analyte by cognitive status (MCI vs CU; Fig. 2A), amyloid PET status (Fig. 2B), tau PET status (Fig. 2C), and synSAA status (Fig. 2D). Detailed summaries of the proteins differentially expressed in each comparison are shown in Supplementary Table 1.

**Figure 2.**
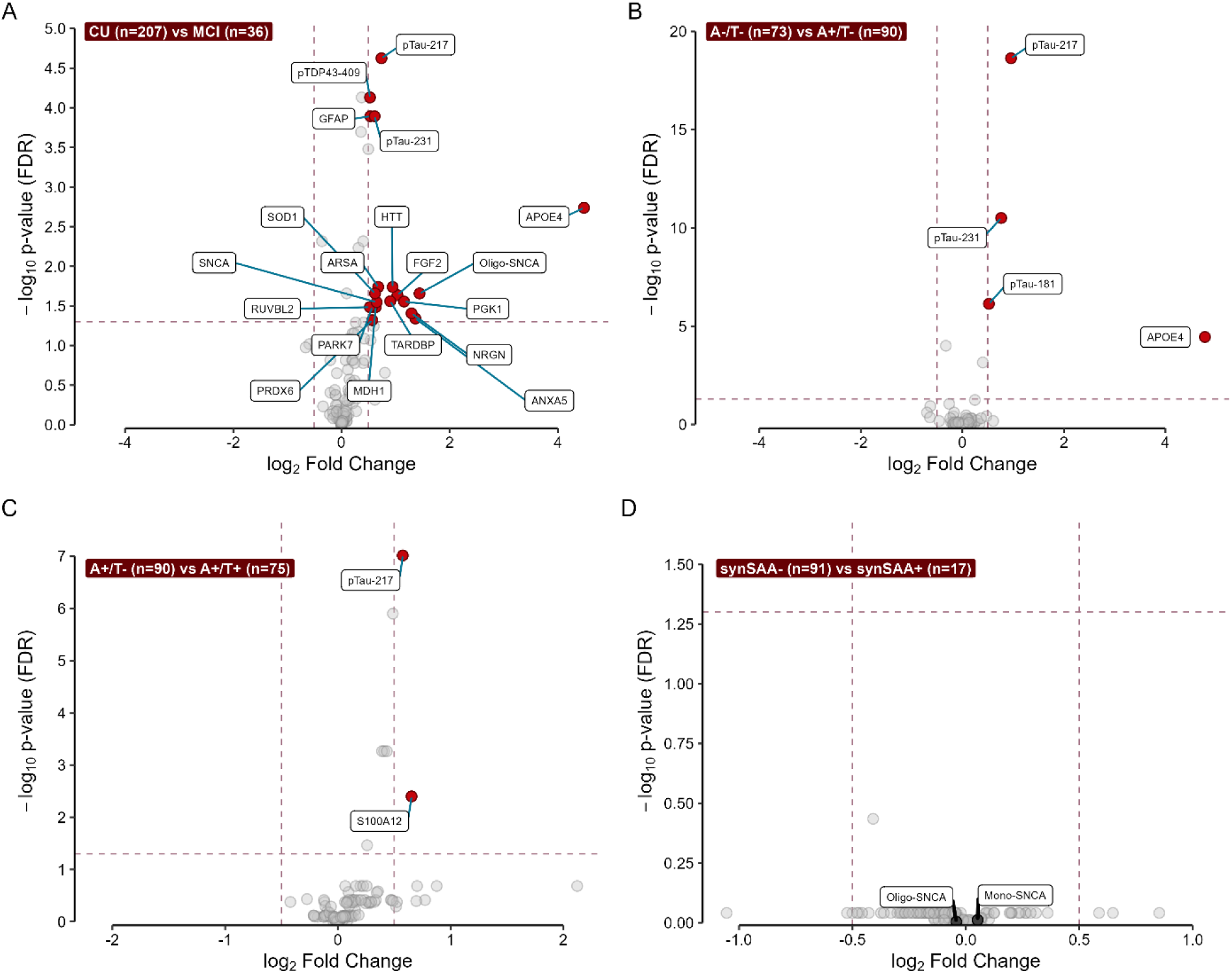
Proteinomics analysis across groups of interest. We performed comparisons between binary groups of interest across all biomarkers in the CNS120 panel. We only considered as analytes of interest those that showed a log_2_ fold change between categories of at least 0.5 (vertical dashed lines), and that maintained a p-value < 0.05 after FDR correction (horizontal dashed line). Analytes meeting these criteria for each of the tests are highlighted in red. We stratified participants according to four different variables: A) Cognitive diagnosis status (CU vs MCI); B) Amyloid status (A-/T- vs A+/T-); C) Tau status (A+/T- vs A+/T+); D) synSAA status (synSAA- vs synSAA+). See Supplementary Table 1 for details about the proteins differentially expressed in each comparison.

We identified 19 proteins that were differentially expressed in participants with a clinical diagnosis of MCI (n=36) compared to those without cognitive impairment (n=207), all of which were upregulated in the MCI group (Figure 2A). Among these were AD-related proteins, including pTau217, pTau231, and APOE4. Interestingly, several candidate biomarkers of other neurodegenerative pathologies were also present. These included pTDP43-409 (log_2_FC = 0.53) and transactive response DNA-binding protein (TARDBP, log_2_FC = 0.9), candidate markers of TDP-43-related diseases such as LATE and FTLD; oligomeric alpha-synuclein (Oligo-SNCA, log_2_FC = 1.44) and monomeric alpha-synuclein (SNCA, log_2_FC = 0.65), potentially indicative of Lewy body dementia; and huntingtin (HTT, log_2_FC = 0.95), related to Huntington’s disease.

Additionally, six proteins were significantly upregulated in participants with a clinical diagnosis of dementia (n=9) compared to those without cognitive impairment (Supplementary Figure S1A). Three of these proteins, neurofilament light polypeptide (NEFL), acetylcholinesterase (ACHE), and pTau181 were uniquely upregulated in participants with dementia and not in MCI participants. No differences in protein expression were found when comparing participants with dementia diagnoses to those with MCI (Supplementary Figure S1B). However, this may be due to the limited number of participants with dementia or MCI in our sample.

To determine differentially expressed proteins due to the presence of amyloid, but independent of tau, we compared A-T- participants (n=73) to the A+T- (n=90) group (Figure 2B). Four proteins were significantly upregulated in A+ participants: APOE4 (log_2_FC = 4.78), pTau217 (log_2_FC = 0.96), pTau231 (log_2_FC = 0.76), and pTau181 (log_2_FC = 0.52). Similarly, to identify proteins related to tau independent of amyloid, we stratified participants into A+T- (n=90) and A+T+ (n=75) groups (Figure 2C). Only two proteins were significantly different in the T+ group, pTau217 and S100 calcium-binding protein A12 (S100A12), both upregulated in T+ participants (log_2_FC = 0.58 and 0.65, respectively). Comparing the extremes of the A/T spectrum, A-/T- and A+/T+ (Supplementary Figure S1C), revealed eight differentially expressed proteins in the diseased group. Among these, four proteins were also upregulated in the A+/T- group: APOE4 (log2FC = 6.9), pTau217 (log_2_FC = 1.53), pTau231 (log_2_FC = 1.2), and pTau181 (log_2_FC = 0.91).

Additional markers included neurofilament heavy polypeptide (NEFH, log_2_FC = 1.49), GFAP (log_2_FC = 0.89), total tau (MAPT, log_2_FC = 0.51), all upregulated in A+T+, and one downregulated marker, C-reactive protein (CRP, log_2_FC = -0.76).

### Biomarkers for Non-AD pathologies

There were no significant differences in biomarker expression between synSAA- (n = 92) and synSAA+ (n = 17) participants based on the volcano plot (all p_FDR_ > .05). APOE4 showed the largest fold-change between the two groups (log2 fold change = 1.47; p_FDR_ = .99). There was considerable overlap between plasma measurements of NULISAseq oligomeric alpha-synuclein (Oligo-SNCA) when participants were stratified by synSAA status (Figure 3A), with no significant distinction between synSAA positive and negative groups based on Oligo-SNCA. Non-pathological monomeric alpha-synuclein (Mono-SNCA, Figure 3B) showed similar trends. It is important to note that the number of participants in this study with known aggregated alpha-synuclein status is relatively small (n = 108) and may not be appropriately powered to detect differences between groups using these biomarkers.

**Figure 3.**
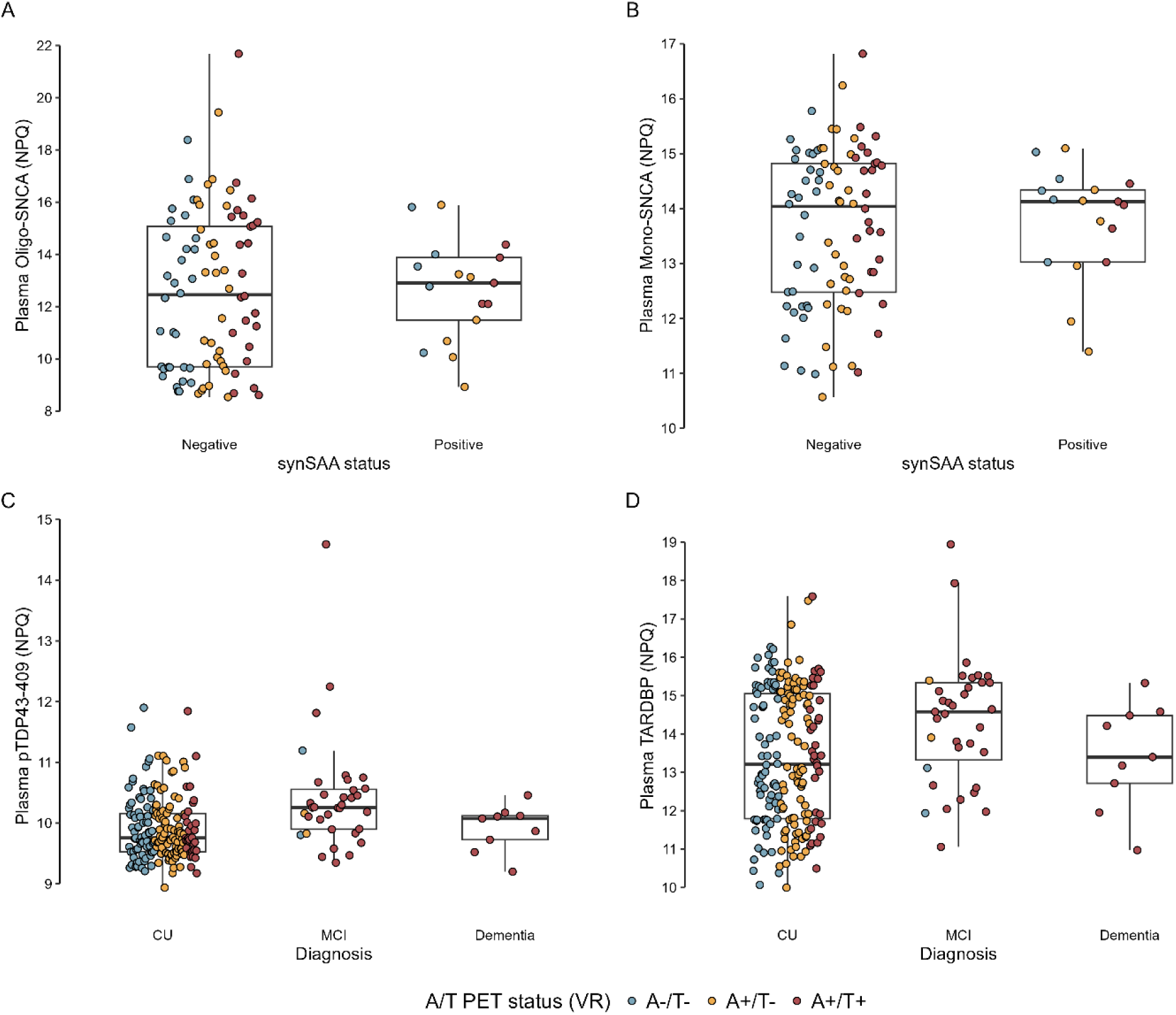
Distribution of non-AD biomarkers by categories of interest. We first compared plasma biomarker expression in participants with results from the CSF Amprion seed amplification assay (synSAA), classified as synSAA- (n =68) and synSAA+(n = 17). A) plasma Oligo-SNCA and B) plasma monomeric alpha-synuclein (Mono-SNCA). Given the results from the proteinomics analysis identifying expression of plasma biomarkers between CU and MCI individuals, we additionally looked at the distribution of plasma pTDP43 (C) and plasma TARDBP (D) among cognitive diagnosis statuses. We additionally show the A/T PET status in distinct colors in all these panels.

### Exploratory Follow-up Analyses

As previously noted, pTDP43-409 was upregulated in participants with MCI compared to those with normal cognition. Based on this observation, we examined the relationship between binarized pTDP43-409 and longitudinal cognition and hippocampal volumes. Using binarized pTDP43-409, we found a significant three-way interaction between A/T status, pTDP43-409 level, and age for both cognitive composites analyzed (PACC3-TMTB: *F*(2, 1170.15) = 7.40, *p* < 0.001; Memory: *F*(2, 1176.80) = 4.30, *p* = 0.014). Estimated simple slopes indicated that participants with both A+/T+ status and elevated levels of pTDP43-409 showed steeper cognitive decline compared to those with Non-elevated pTDP43-409 or A-/T- Figure 4A and 4B).

**Figure 4.**
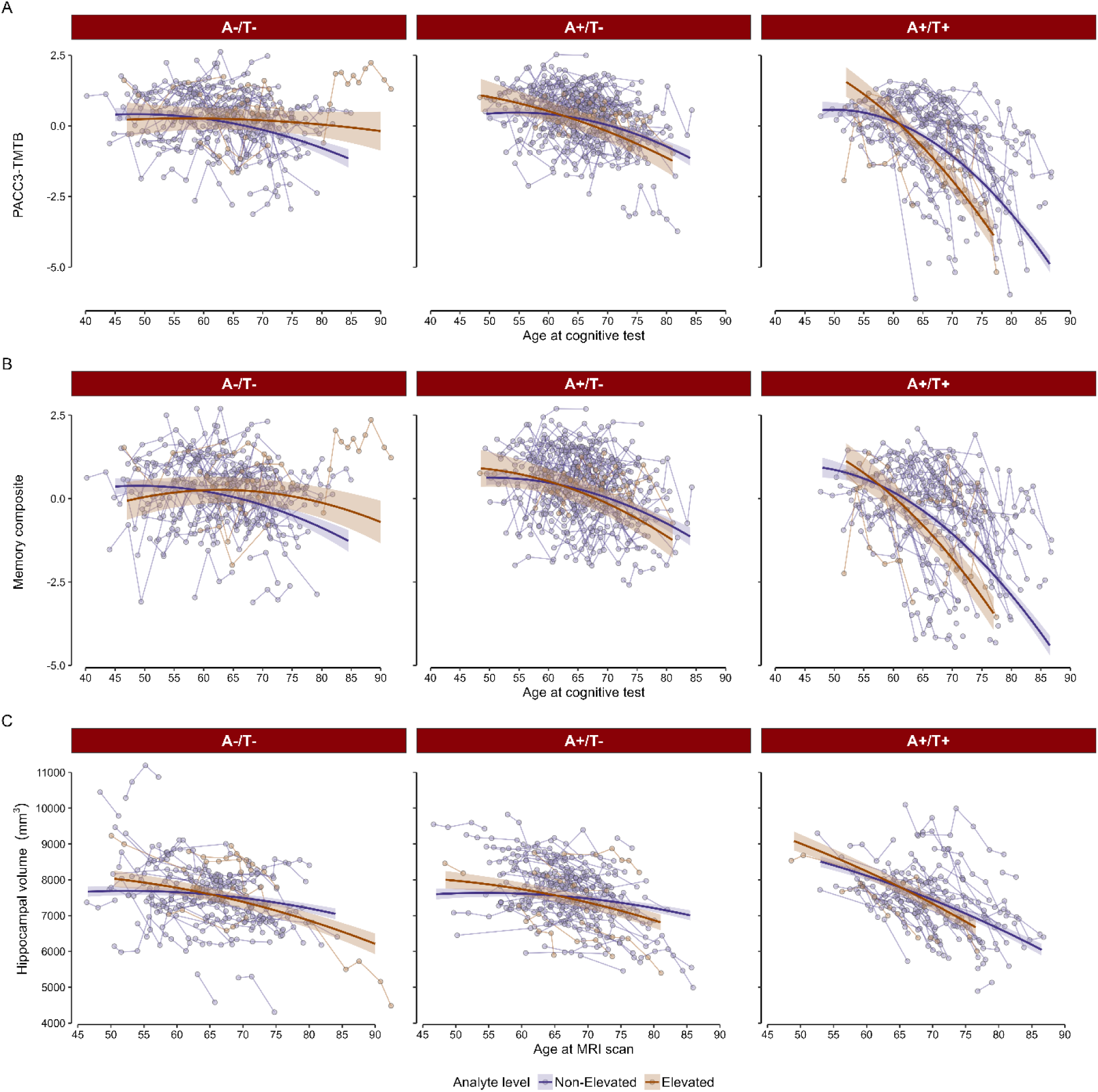
Estimated and observed longitudinal trajectories among pTDP43 groups. A-B) Predicted slopes across time in each of the pTDP43 groups depending on their A/T status, for the PACC3-TMTB and memory composite scores, respectively. C) Predicted slopes for hippocampal volume across time in each of the pTDP43 groups depending on their A/T status. Solid dark lines represent the marginal effects across time for each pTDP43 group, shaded areas correspond to the SE. Dots and lines in the background represent the observed values from everyone in the sample.

After sequentially removing higher order non-significant interactions, the pTDP43-409 group by age interaction was significant in regard to hippocampal atrophy (F(1, 839.31) = 7.93, *p* = .005) as was the PET status by age interaction (*F*(2, 823.23) = 35.42, *p* < .001). Full final model output, shown in Table 2, and simple slopes, shown in Figure 4C, depict the faster decline in hippocampal volume in those who have elevated pTDP43-409 and/or A+/T+.

## Discussion

In this study, we evaluated the performance and potential utility of the NULISAseq CNS panel for characterizing AD pathology and detecting non-AD proteinopathies. We found that the NULISAseq CNS panel identified several known markers of AD that are upregulated in amyloid positive participants, most importantly pTau217, and that the performance of pTau217 as part of the panel was comparable to Simoa in differentiating amyloid PET status. These findings are in line with other early studies utilizing the NULISAseq CNS panel [32–36] and aid in validating use of this panel for detecting AD. This validation is critical for investigating other proteinopathies that often co-occur with AD since most neurodegenerative cohorts are AD-focused and biomarkers for other neurodegenerative dementias are not widespread. Thus, anchoring exploratory biomarker trends to established AD biomarkers provides some context to early-stage studies such as ours.

The pTau217-NULISA and pTau217-Simoa assays (both using ALZPath antibodies) were not significantly different in terms of their ability to differentiate amyloid PET status in this sample. The AUC from pTau217-NULISA (0.92, 0.88-0.97) is similar to those reported in recent studies using other pTau217 assays and independent cohorts [37, 38], indicating consistent performance with other platforms, and is also similar to recent comparisons between NULISAseq pTau217 to PET amyloid status [34] or CSF [39] in other cohorts.

Both the NULISAseq and Simoa pTau217 assays accurately predict tau PET status with ROC AUCs exceeding 0.80. Slightly lower performance in tau detection compared to amyloid is not unexpected given that pTau217 is a Core 2 marker in relation to tau *staging* whereas it is a Core 1 marker for amyloid *detection* [1]. This distinction is important and likely due to the heterogeneity of manifested tau burden following amyloid onset. Notably, the Simoa pTau217 assay was slightly better at classifying tau status than NULISAseq pTau217 despite both using the same antibody. Differences in performance may be due to assay platforms as well as NULISAseq pTau217 being part of a large multiplex panel, which requires a wide dynamic range to detect both high and low abundance proteins, whereas Simoa pTau217 is a single plex. Interestingly, others have reported higher fold-changes for the NULISA pTau217 single plex assay compared to NULISAseq pTau217, though differences were not significant [32].

Our analysis of differential protein expression in A-/T- vs A+/T- showed that A+ participants presented elevated levels of biomarkers in addition to pTau217 that were previously reported in other studies, specifically pTau181 [40], pTau231 [41], GFAP [42], and APOE4 [43]. The increase in APOE4 levels in A+ individuals is expected and driven by the increased prevalence of the AD susceptibility gene *APOE* ε4 in this cohort. Interestingly, our results also showed elevated levels of a calcium-binding protein, S100A12, in A+/T+ relative to A+/T- participants. While relationships between S100 proteins have been observed with AD neuroinflammation [44] and vascular inflammation processes [45], the specific role of these proteins in AD remains unclear.

Currently, our field’s focus is shifting towards the development of biomarker panels to aid in AD staging and detecting other markers of neurodegeneration for co-occurring pathologies [1, 46]. Our results from the differential protein expression in CU vs MCI participants showcase the importance of creating these biomarker profiles to better characterize multi-etiology dementia. Besides biomarkers for amyloid positivity (pTau217, pTau231, GFAP, APOE4), we found elevated plasma levels of proteins related to inflammatory and oxidative stress processes in neurodegeneration (e.g., peroxiredoxin 6 [47], malate dehydrogenase 1 [48], superoxide dismutase 1 [49]), proteins related to LATE (TARDBP and pTDP43-409 [50]), proteins related to dementia with Lewy bodies and Parkinson’s disease (alpha synuclein), and Huntington’s disease (huntingtin [51]), among others. These now require further validation and replication in larger samples as it is likely that some of the signal we observed was related to person-level heterogeneity in pathology burden.

One of the advantages of a multiplexed biomarker panel is the potential to detect non-AD neurodegenerative pathologies. The presence of these pathologies along with AD may contribute to heterogeneity in clinical symptoms, disease progression, and treatment response. Alternatively, if a non-AD pathology is the primary driver of symptoms, excluding AD is necessary to guide treatment, especially in the era of disease-modifying AD therapeutics. The NULISAseq CNS panel contains multiple biomarkers implicated in non-AD pathologies, but their performance is relatively untested in larger cohort studies or neuropathology-based studies. For example, a recent study reported in medRxiv also using the NULISAseq CNS panel found upregulated proteins related to TDP-43 and alpha-synuclein pathologies, both of which commonly co-occur with AD [5], in a cohort of 20 progranulin (GRN) mutation carriers compared to matched controls [32]. Since pTDP43-409 and Oligo-SNCA were also upregulated in our impaired group, we further explored the relationship between these markers and other available fluid, imaging, and cognitive data.

Our analyses with alpha-synuclein biomarkers did not show differential expression of any proteins in our sample between participants with synSAA+ and synSAA-. Although the number of positive observations in this subset of our sample is small, this suggests that in plasma, the antibody used in the NULISAseq Oligo-SNCA assay cross-reacts with other, perhaps more abundant, species of SNCA, such as the monomeric, non-pathological form [52]. Similar findings were reported in [32].

In the pTDP43-409 analysis, we observed faster decline in both hippocampal volume and cognitive scores in those participants with higher concentrations of this protein in plasma, compared to those with lower concentrations. These findings demonstrate the potential utility of the NULISAseq platform for identifying non-AD co-occurring pathology. However, these preliminary results require validation through autopsy-confirmed TDP-43 cases. This work is currently in progress.

Our study was intended as proof-of-concept, focusing first on identifying known AD biomarkers of participants with well-characterized biomarker profiles using previously validated methods [37], and then detecting additional biomarkers indicative of other underlying conditions. The finding of upregulated pTDP43-409 in some participants from the MCI group was unanticipated though plausible based on the follow up analyses showing associations of pTDP43-409 with cognition and hippocampal atrophy.

A limitation of this study is that the samples utilized here were from cohorts focused on AD detection and characterization. Thus, in-depth clinical phenotyping of non-AD dementias was not available for these participants. Additionally, the current study only classified participants as T+ if they had signal in both medial temporal lobe and neocortex, therefore we have some heterogeneity in the A+T- group, which includes some participants with tau signal in the medial temporal lobe (n = 14) and some with signal in the neocortex (n = 9). Recent work has found that participants with A+ status and significant tau PET signal in either the medial temporal lobe or neocortex can present elevated pTau217 levels and significant cognitive decline compared to those without it [20], which would be relevant to consider in future work.

We stress that our exploratory analyses are preliminary and need further confirmation and delineation of a clear threshold based on gold standard autopsy cases to indicate elevated vs normal pTDP43-409. Further, the small sample size in this preliminary study is a limitation and an expanded study is underway based on these encouraging early results. It will be important for future studies to investigate longitudinal samples on the panel of biomarkers with particular attention on the AD and putative LATE markers.

In conclusion, these preliminary results demonstrate the advantage of highly multiplexed assays in characterizing amyloid and tau pathology and the role of other markers on the panel that may inform on other pathologies or on the severity of neurodegeneration and other tissue reactions. However, further investigation is needed, including replication in other cohorts. We were particularly intrigued by the pTDP43-409 findings with both cognitive decline and hippocampal neurodegeneration suggesting that pTDP43-409 could be a promising blood biomarker for detecting TDP-43 clinical syndrome and perhaps the proteinopathy. Further studies in longitudinal, well-characterized cohorts are needed to determine patterns of protein expression relative to symptom onset, presentation, and therapeutic outcomes.

## Data Availability

All data produced in the present study are available upon reasonable request to the authors

https://wrap.wisc.edu/data-requests-2/

## Acknowledgements

The authors would like to thank WRAP and WADRC participants and staff for their contributions. Without their efforts, this research would not be possible. HZ is a Wallenberg Scholar and a Distinguished Professor at the Swedish Research Council supported by grants from the Swedish Research Council (#2023-00356; #2022-01018 and #2019-02397), the European Union’s Horizon Europe research and innovation program under grant agreement No 101053962, Swedish State Support for Clinical Research (#ALFGBG-71320), the Alzheimer Drug Discovery Foundation (ADDF), USA (#201809-2016862), the AD Strategic Fund and the Alzheimer’s Association (#ADSF-21-831376-C, #ADSF-21-831381-C, #ADSF-21-831377-C, and #ADSF-24-1284328-C), the European Partnership on Metrology, co-financed from the European Union’s Horizon Europe Research and Innovation Programme and by the Participating States (NEuroBioStand, #22HLT07), the Bluefield Project, Cure Alzheimer’s Fund, the Olav Thon Foundation, the Erling-Persson Family Foundation, Familjen Rönströms Stiftelse, Stiftelsen för Gamla Tjänarinnor, Hjärnfonden, Sweden (#FO2022-0270), the European Union’s Horizon 2020 research and innovation programme under the Marie Skłodowska-Curie grant agreement No 860197 (MIRIADE), the European Union Joint Programme – Neurodegenerative Disease Research (JPND2021-00694), the National Institute for Health and Care Research University College London Hospitals Biomedical Research Centre, and the UK Dementia Research Institute at UCL (UKDRI-1003).

## Consent statement

The study procedures received approval from the University of Wisconsin-Madison Institutional Review Board and were conducted in compliance with the World Medical Association Declaration of Helsinki. All subjects provided informed consent.

