## Supplementary Materials for "Targeted Proteomic Biomarker Profiling Using NULISA in a cohort enriched with risk for Alzheimer’s Disease and Related Dementias"

Supplementary Table 1

| **Protein name** | **Abbreviation** | **log_2_ Fold Change** | ***p_FDR_* value** |
| --- | --- | --- | --- |
| ***CU vs MCI*** | | | |
| Apolipoprotein E isoform 4 | APOE4 | 4.48 | = .002 |
| Oligomeric alpha-synuclein | Oligo-SNCA | 1.44 | = .022 |
| Annexin A5 | ANXA5 | 1.37 | = .046 |
| Neurogranin | NRGN | 1.3 | = .039 |
| Phosphoglycerate kinase 1 | PGK1 | 1.16 | = .028 |
| Fibroblast growth factor 2 | FGF2 | 1.04 | = .023 |
| Huntingtin | HTT | 0.95 | = .018 |
| TAR DNA-binding protein 43 | TARDBP | 0.9 | = .028 |
| Phosphorylated tau-217 | pTau-217 | 0.74 | < .001 |
| Arylsulfatase A | ARSA | 0.68 | = .018 |
| Monomeric alpha-synuclein | SNCA | 0.65 | = .028 |
| Malate dehydrogenase, cytoplasmic | MDH1 | 0.63 | = .033 |
| Superoxide dismutase [Cu-Zn] | SOD1 | 0.62 | = .022 |
| Phosphorylated tau-231 | pTau-231 | 0.61 | < .001 |
| Parkinson disease protein 7 | PARK7 | 0.57 | = .033 |
| Peroxiredoxin-6 | PRDX6 | 0.57 | = .048 |
| TDP-43 with phosphorylation on serine 409 | pTDP43-409 | 0.53 | < .001 |
| Glial fibrillary acidic protein | GFAP | 0.53 | < .001 |
| RuvB-like 2 | RUVBL2 | 0.52 | = .033 |
| ***A-/T- vs A+/T-*** | | | |
| Apolipoprotein E isoform 4 | APOE4 | 4.78 | < .001 |
| Phosphorylated tau-217 | pTau-217 | 0.96 | < .001 |
| Phosphorylated tau-231 | pTau-231 | 0.76 | < .001 |
| Phosphorylated tau-181 | pTau-181 | 0.52 | < .001 |
| ***A+/T- vs A+/T+*** | | | |
| S100 Calcium Binding Protein A12 | S100A12 | 0.65 | = .004 |
| Phosphorylated tau-217 | pTau-217 | 0.58 | < .001 |
| ***A-/T- vs A+/T+*** | | | |
| Apolipoprotein E isoform 4 | APOE4 | 6.9 | < .001 |
| Phosphorylated tau-217 | pTau-217 | 1.53 | < .001 |
| Neurofilament heavy polypeptide | NEFH | 1.48 | = .005 |
| Phosphorylated tau-231 | pTau-231 | 1.2 | < .001 |
| Phosphorylated tau-181 | pTau-181 | 0.91 | < .001 |
| Glial fibrillary acidic protein | GFAP | 0.89 | < .001 |
| Microtubule-associated protein tau (total tau) | MAPT | 0.51 | < .001 |
| C-reactive protein | CRP | -0.76 | = .019 |
| ***CU vs Dementia*** | | | |
| Phosphorylated tau-217 | pTau-217 | 1.19 | < .001 |
| Phosphorylated tau-231 | pTau-231 | 0.89 | = .018 |
| Acetylcholinesterase | ACHE | 0.86 | < .001 |
| Glial fibrillary acidic protein | GFAP | 0.85 | = .006 |
| Neurofilament light polypeptide | NEFL | 0.69 | = .038 |
| Phosphorylated tau-181 | pTau-181 | 0.69 | = .038 |

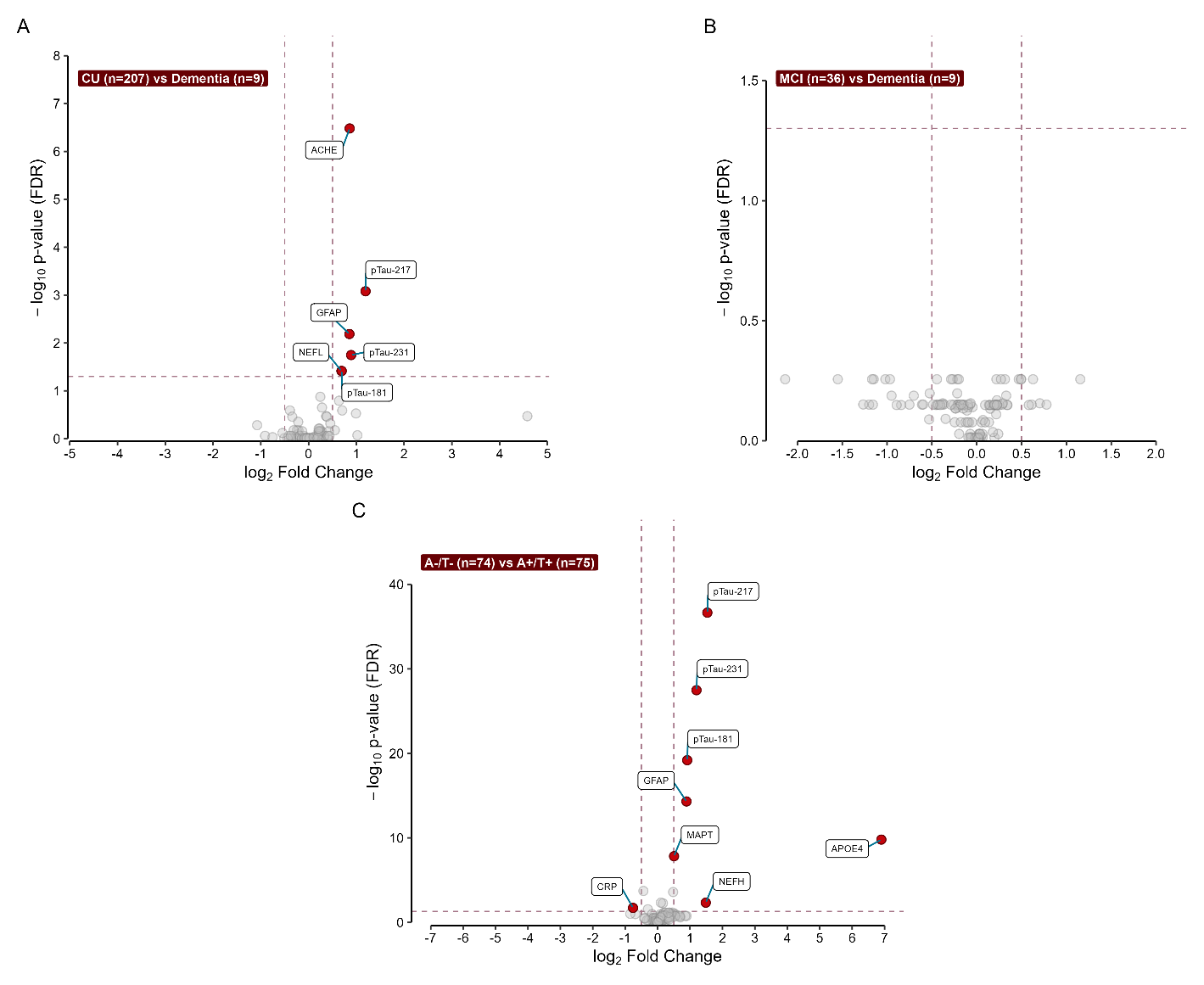

Figure S1. Additional proteomics analysis. We performed comparisons between binary groups of interest across all biomarkers in the CNS120 panel. We only considered as analytes of interest those that showed a log_2_ fold change between categories of at least 0.5 (vertical dashed lines), and that maintained a p-value < 0.05 after FDR correction (horizontal dashed line). Analytes meeting these criteria for each of the tests are highlighted in red. In panels A-B, we stratified participants according to their cognitive status: A) CU vs Dementia; B) MCI vs Dementia. In panel C we show the comparison between A-T- and A+T+.
